# Healthcare Big Data Platform for Linking National Databases in Korea: System Development and Research Applications

**DOI:** 10.64898/2026.07.09.26357705

**Authors:** Yujin Kim, Yerim Lee, Jipmin Jeong

## Abstract

Public healthcare databases in South Korea have been distributed across disparate government agencies, requiring researchers to navigate multiple, separate institutional approval processes for data linkage. To address the systemic inefficiency, the Healthcare Big Data Linkage Platform (HCDL), jointly administered by NECA and KHIS, was established to integrate 13 databases from 10 public institutions through a Trusted Third Party (TTP)-based linkage methodology and a centralized one-stop review process. Of 311 projects submitted between 2022 and 2025, 190 (61.1%) were approved with annual applications increasing 2.4-fold over the study period. The average number of databases per project exceeded three, reflecting a surging demand for integrated clinical data. Nationwide healthcare data from HIRA and NHIS were the most frequently requested databases (93.2% and 82.1% of approved projects, respectively), and co-occurrence pattern analysis further confirmed that both formed the core of the research ecosystem in combination with vital status, lifestyle, and cancer diagnosis data. By consolidating multi-institutional review and enabling equitable data access, the HCDL has emerged as a core infrastructure for data-driven and precision medicine research in South Korea.

**Background:** Public healthcare databases in South Korea have been managed in a fragmented manner across disparate government agencies, requiring researchers to contact each institution individually and complete separate approval processes. This structural inefficiency significantly prolonged the time from data request to use and constrained research to datasets available within a researcher’s own institutional network.

**Objective:** This study describes the system design and governance structure of the Healthcare Big Data Linkage Platform (HCDL) and examines data utilization trends from 2022 to 2025 to evaluate the platform’s impact on the domestic healthcare research environment.

**Methods:** The HCDL, jointly administered by the National Evidence-based Healthcare Collaborating Agency (NECA) and the Korea Health Information Service (KHIS), integrates 13 databases from 10 public institutions. The platform employs a Trusted Third Party (TTP)-based linkage methodology and a centralized Research Evaluation Committee (REC) for one-stop review. Data on application submissions, approvals, database usage patterns, and inter-database co-occurrence networks across 311 submitted projects were analyzed.

**Results:** A total of 311 projects were submitted between 2022 and 2025, of which 190 (61.1%) were approved. Annual applications increased approximately 2.4-fold, from 44 in 2022 to 105 in 2025, with approval rates remaining stable between 51.7% and 66.7%. CLAIMS and SCREEN were the most frequently requested databases (93.2% and 82.1% of approved projects, respectively), suggesting strong and consistent demand for nationwide healthcare data as the analytical foundation for research. Co-occurrence pattern analysis further revealed that the CLAIMS-SCREEN pairing formed the core of the research ecosystem in combination with vital status, lifestyle, and cancer diagnosis data.

**Conclusions:** The HCDL has substantially reduced administrative burden and expanded data accessibility by consolidating multi-institutional review and linkage into a single integrated process. By enabling equitable access to public health, the platform has emerged as a core infrastructure supporting the growth of data-driven and precision medicine research in South Korea. Further development priorities include expanding analytical capacity, incorporating clinical data sources such as electronic medical records, and establishing a standardized metadata framework to enhance interoperability.

## INTRODUCTION

Healthcare data represent a broad resource encompassing all information related to physical and mental health, including individual health status, disease, nutrition, and welfare (Musa et al., 2023). In Korea, healthcare data are generated and managed primarily by medical institutions and public agencies. Healthcare providers generate large volumes of data such as medical records, prescriptions, and test results, during patient care, which are owned and managed within each institution. Public agencies, by contrast, collect and manage comprehensive population-level datasets for the purposes of national health policy development and public health research. These include health insurance eligibility and premium records, health examination results, and healthcare utilization data managed by the National Health Insurance Service (NHIS) and the Health Insurance Review and Assessment Service (HIRA) (Kang et al., 2018), as well as communicable disease surveillance data (Park & Cho, 2014) and the Korea National Health and Nutrition Examination Survey (KNHANES) (Oh et al., 2021; Kang et al., 2018) maintained by the Korea Disease Control and Prevention Agency (KDCA). These public healthcare datasets are essential resources for national health policy development, epidemiological research, and healthcare service evaluation (Jang et al., 2025).

However, these critical data resources have historically been stored in a fragmented manner across multiple government agencies, limiting data accessibility and usability (Kim et al., 2017). Under the conventional approach through dedicated linkage agency, researchers seeking to link data from multiple institutions were required to contact each agency individually, complete separate and often heterogeneous approval processes sequentially for each institution, coordinate and await data linkage schedules on an institution-by-institution basis, independently verify and negotiate the level of pseudonymization applied to linked data, and submit separate requests for data export approvals. This fragmented and non-standardized process significantly prolonged the period from data request to actual use (Lee et al., 2024), and has been widely identified as a structural impediment to advancing healthcare research (Figure 1).

**Figure 1.**
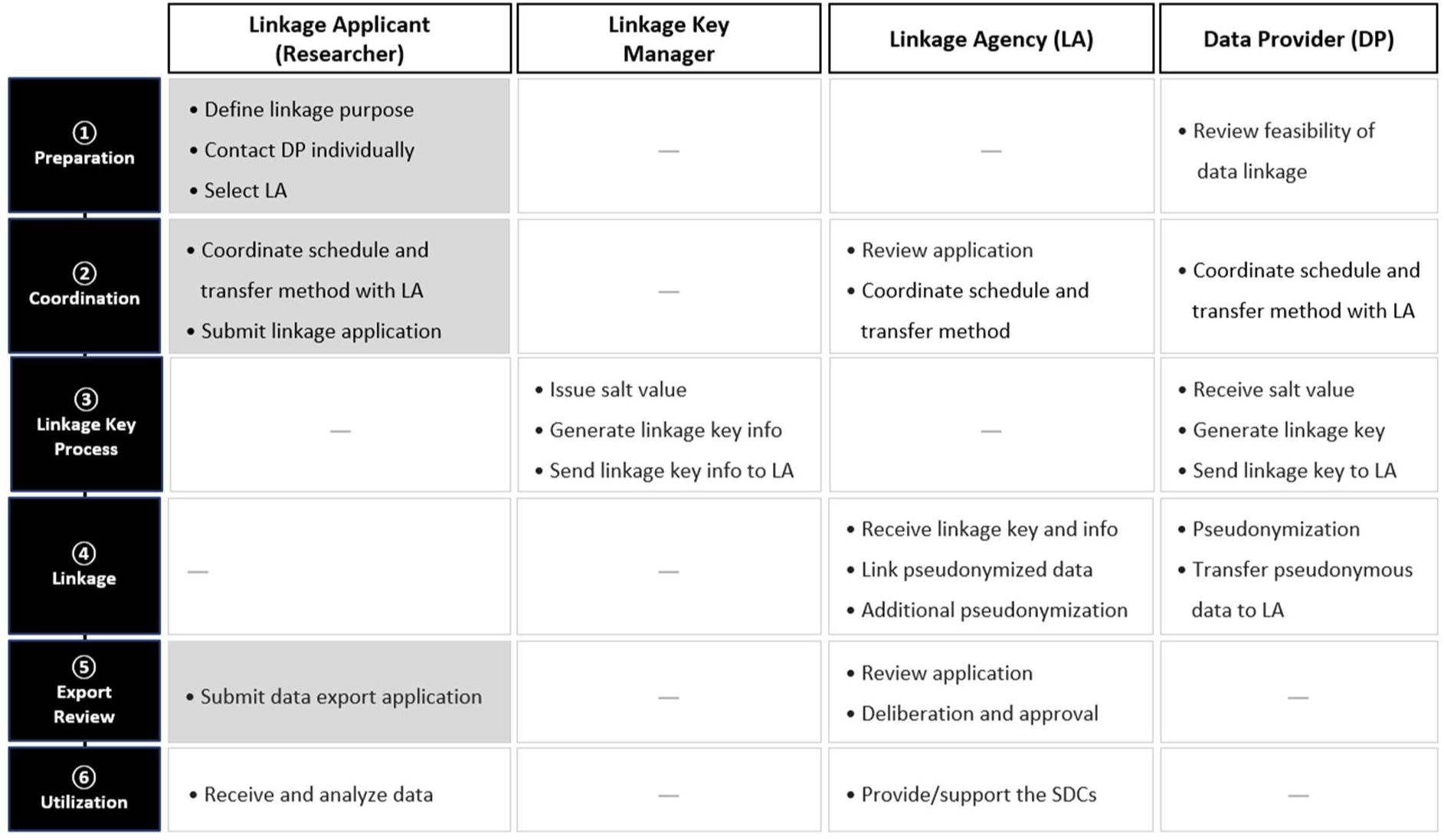
Conventional data linkage procedure via linkage agency.

Furthermore, conventional data linkage by a specialized agency imposed practical limitations on the scope of accessible data. Although researchers were not categorically prohibited from linking data held by institutions outside their own affiliation, data-holding agencies in practice tended to be reluctant to share data with external researchers, making inter-institutional negotiations difficult. As a result, researchers were effectively constrained to linking data held by their own institution or institutions with which they had established collaborative relationships. This constituted a substantive barrier that limited the diversity and depth of national healthcare research (Jang et al., 2025).

A number of countries have developed integrated platforms to overcome these limitations. Finland established Findata (the Finnish Health and Social Data Permit Authority) in 2020 under the Act on the Secondary Use of Health and Social Data, providing a single-window application system and secure remote analysis services (Kapseli) for data distributed across multiple agencies, including the Finnish Institute for Health and Welfare (THL), the Social Insurance Institution of Finland (Kela), and Statistics Finland (Vilpponen et al., 2024). Estonia leverages its national digital infrastructure, X-Road, to interlink nationwide EMRs with national registry data, providing the foundation for large-scale genomic-clinical linkage research through the Estonian Biobank operated by the University of Tartu (Leitsalu et al., 2015). In the United Kingdom, the Clinical Practice Research Datalink (CPRD) links general practitioner (GP) records with hospital inpatient data (Hospital Episode Statistics; HES), mortality records (Office for National Statistics; ONS), and cancer registry data (National Cancer Registration and Analysis Service; NCRAS), with research validity assessed through review by an Independent Scientific Advisory Committee (ISAC) (Herrett et al., 2015). These countries share the characteristic that population-wide data underpinned by universal insurance or public healthcare systems are centralized within a single infrastructure.

In South Korea, the Healthcare Data Linkage Platform (HCDL) was developed under the leadership of the Ministry of Health and Welfare (MOHW) to address these challenges and more effectively support data use for research. The platform has two key features distinguished from conventional approaches. First, researchers no longer need to contact individual data-holding agencies; instead, they can identify the data they need through standardized data specifications provided by the platform and submit a single application through the platform. All subsequent steps such as institutional review, data linkage, pseudonymization, and export review, are processed in an integrated manner through the platform, substantially reducing the administrative burden on researchers. Second, researchers can combine datasets from all participating public institutions regardless of their institutional affiliation. This expands the scope and combinatorial possibilities of healthcare research, which would otherwise be structurally infeasible under the conventional linkage agency system.

The HCDL was initiated as a policy research project in 2015, followed by pilot operations from 2018 to 2022, with full-scale operations beginning in 2023. The platform is jointly administered by the National Evidence-based Healthcare Collaborating Agency (NECA) and the Korea Health Information Service (KHIS). This study examines the platform’s background, governance structure, TTP-based data linkage methodology, pseudonymization framework, and data use procedures. It also reviews the data utilization trends from 2022 to 2025, with the aim of elucidating the structural transformation that the platform has brought to the domestic healthcare research environment and its broader implications.

## METHODS

### Participating institutions and Available Datasets

Nine public institutions and one national hospital currently participate in the platform, collectively providing 13 databases. The Korea Disease Control and Prevention Agency (KDCA) contributes five databases, while the remaining institutions each maintain separate databases designated for platform use. The dataset composition is summarized in Table 1.

**Table 1.** List of Available Datasets (as of December 2025)

| Institution | Coverage Years | Dataset Name | Acronym |
| --- | --- | --- | --- |
| Korea Disease Control and Prevention Agency (KDCA) | '07-'22 | KNHANES Basic DB | KNHANES |
|  | '07-'23 | KNHANES Air Pollution DB | AIR |
|  | '13-'24 | Tuberculosis Notification DB | TUBER |
|  | '01-'13 | Korean Genome and Epidemiology Study | KoGES |
|  | '12-'24 | National Immunization DB | IMM |
| Ministry of Data and Statistics (MODS) | '12-'22 | Cause of Death Statistics | MORT |
| Military Manpower Administration (MMA) | '15-'24 | Physical Examination Items and Military Draft Screening Results | MIL |
| National Institute of Organ, Tissue and Blood Management (NIOTBM) | '12-'23 | Donor and Recipient Information | DON |
| National Rehabilitation Center (NRC) | '12-'22 | Nursing Clinical Observation Records and Medical Record Information | REHAB |
| National Medical Center (NMC) | '18-'23 | Dementia Screening, Diagnosis, Differential Testing, Case Management, and Dementia Program Data | DEM |
| Health Insurance Review and Assessment Service (HIRA) | '12-'22 | Medical Billing Statements and Healthcare Utilization Records | CLAIMS |
| National Cancer Center (NCC) | '99-'22 | Cancer Registry Data | CANCER |
| National Health | '13-'23 | Eligibility, Insurance Premiums, General | SCREEN |
| Insurance Service (NHIS) |  | Health Screenings, and Other NHIS Enrollee Information |  |
| NHIS Ilsan Hospital | '12-'23 | Patient Information, Visits, Diagnoses, Tests (Imaging, Pathology), Surgery, Radiology, and Vital Signs | ILSAN |

### Data Governance

The platform operates working-level consultative meetings with the participating institutions two to four times annually to discuss operational issues. Annual data updates ensure data currency. A centralized Research Evaluation Committee (REC) conducts one-stop reviews of research validity, public interest, and data export appropriateness, simplifying procedures and improving predictability compared to previously separate, institution-specific reviews.

### Data Use Procedure

The data use process consists of six sequential steps, from researcher application to data access, with a total turnaround of approximately six months. Researchers may request data from up to five databases across a maximum of four institutions per project.

The procedure is as follows: ① Submission and Pre-screening: Researchers submit data request documents including a research proposal, and the secretariat reviews whether the application meets the data provision criteria. ② REC review: Expert panelists from academia and industry, civil society representatives, and data-providing institutions jointly assess the scientific validity, public value, and public-interest contribution of the proposed research design, thereby reflecting social consensus on data utilization. ③ Data Linkage and Pseudonymization: Linkage keys and data are combined to generate an integrated dataset, and additional pseudonymization is applied. ④ Export Appropriateness Review: The Export Review Committee conducts a final review of the linked dataset. ⑤ Data Export: The approved dataset is made available to researchers through a closed Secure Data Center (SDC) (Figure 2).

**Figure 2.**
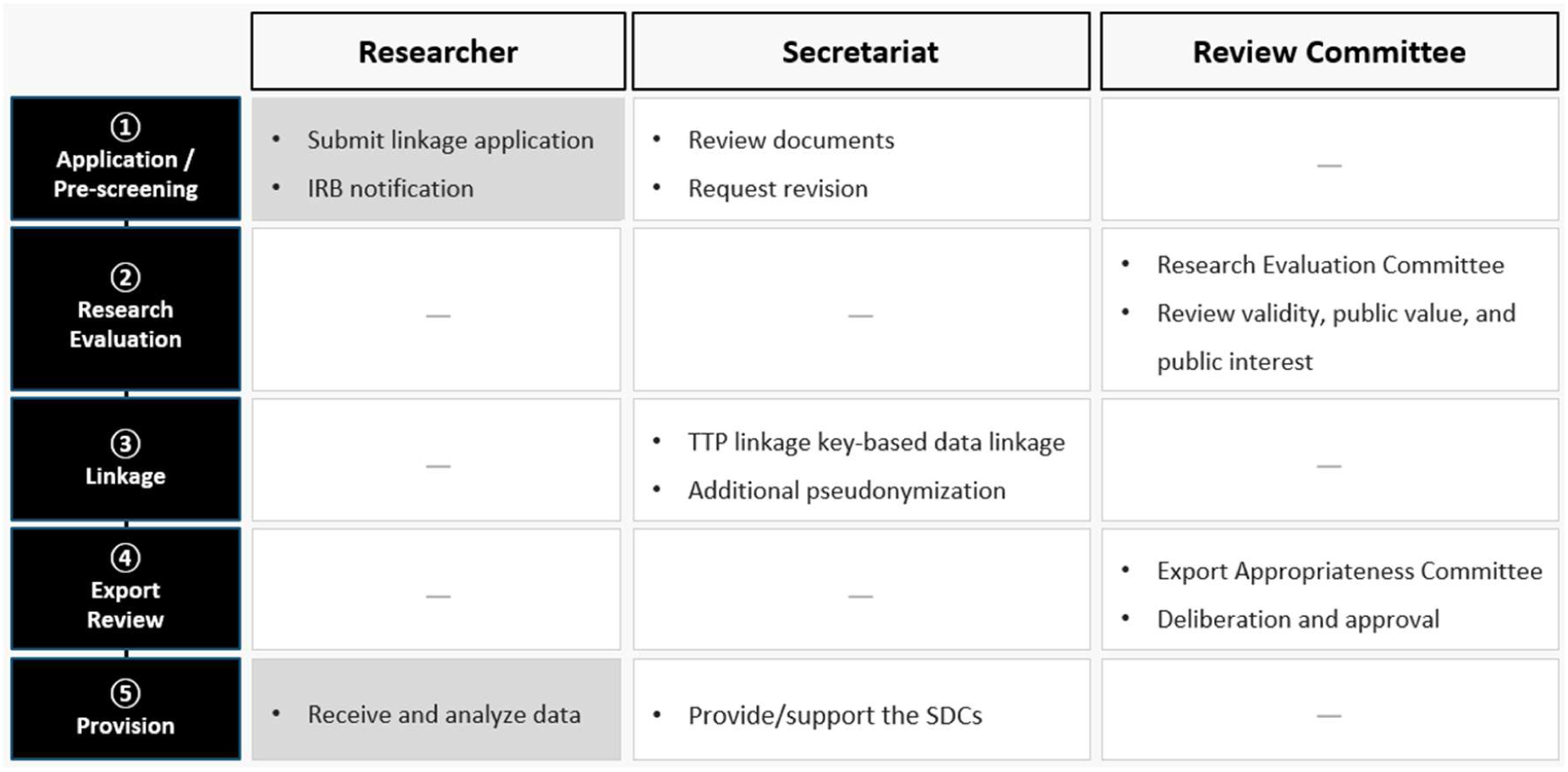
Data use procedure in the HCDL.

### Platform Access

Researchers may apply for data use through the official platform website (https://hcdl.mohw.go.kr) during three to four designated application periods per year. Submission requires a research proposal, data catalog, and Institutional Review Board (IRB) approval, and additional documents such as application for specific disease codes or drug utilization application. Data analysis is restricted to secure, closed-network data centers (SDCs) operated by KHIS, NHIS, HIRA, MODS, and NCC. The standard analysis period is six months, with a one-time extension of up to six months available upon request.

### Data Linkage

Data linkage is performed using a Trusted Third Party (TTP) based key. The Ministry of Health and Welfare issues a 32-character encrypted TTP key to each data-providing institution. Institutions then generate linkage keys by applying a hash function on the TTP key in combination with name, date of birth, and sex. The secretariat consolidates the institution-specific linkage keys into a unified linkage key, which is then used to extract individual-level data from each institution. Upon receipt, the secretariat integrates the data and applies additional pseudonymization (Figure 3).

**Figure 3.**
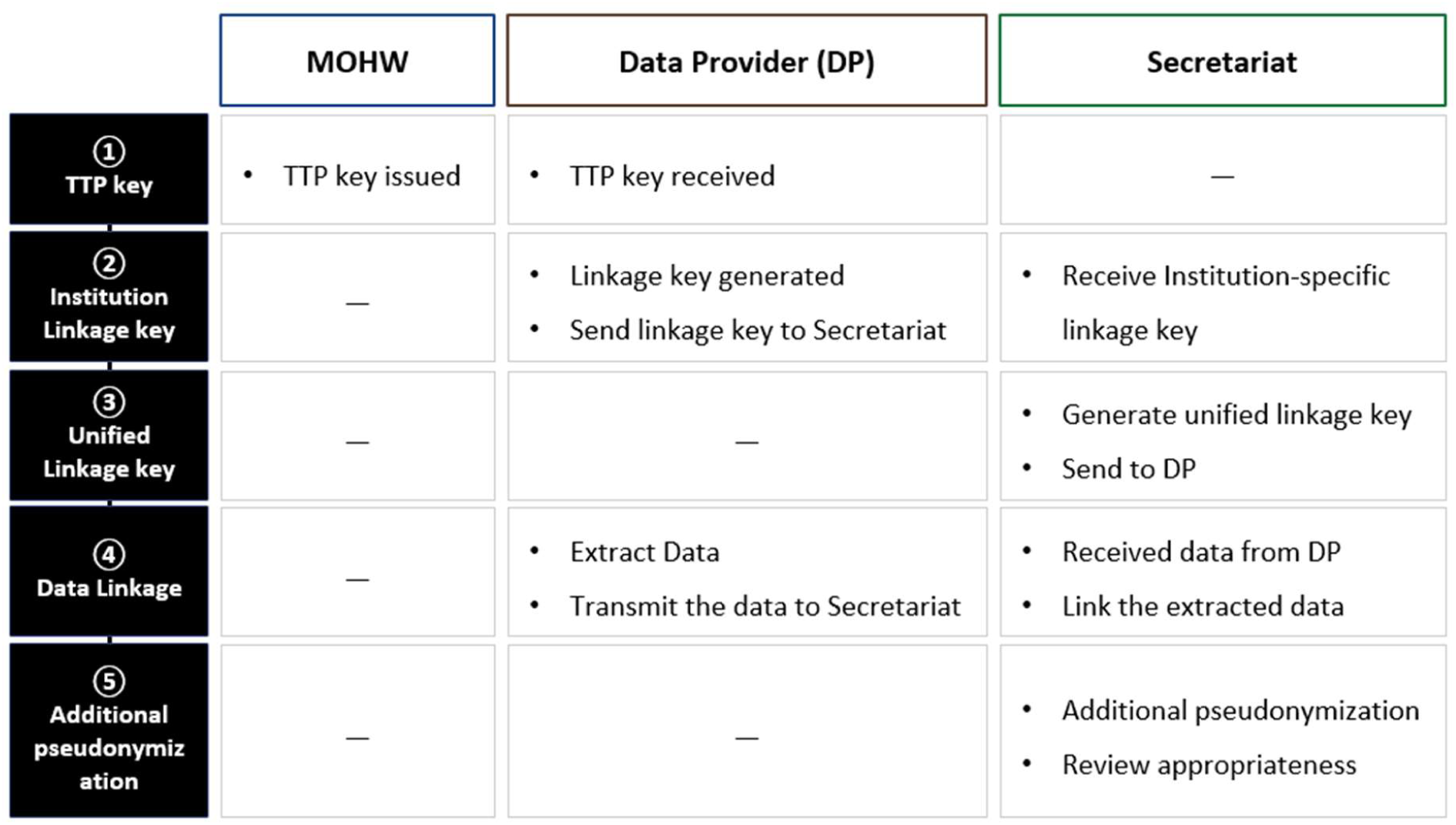
TTP-based data linkage procedure Data Pseudonymization.

The platform performs additional pseudonymization after receiving pre-pseudonymized data from each institution to minimize the risk of re-identification, following the Personal Information Protection Act in Korea.

For quasi-identifiers, address information is provided at the city/county/district (si/gun/gu) level; when sensitive diseases are included, address data are suppressed or aggregated to the provincial level based on annual patient counts. Healthcare provider identifiers are hashed, age is provided in five-year intervals with top-coding applied at 80 years and above, and household size is recategorized into five groups. For general information, dates are provided at the daily level with added random noise (±5 days) while maintaining longitudinal consistency. Drug codes are partially masked, and 493 sensitive disease codes are either suppressed or masked unless separately approved (Table 2).

**Table 2.** Pseudonymization Methods for Sensitive Information.

| Attribute Category | Classification | Pseudonymization Method |
| --- | --- | --- |
| Quasi-identifier | Address | Provided at the city/county/district (si/gun/gu) level within ranges designated by data-holding institutions; when sensitive diseases are included, data are suppressed or aggregated to the provincial level. |
|  | Healthcare provider | Healthcare provider identifiers are hashed |
|  | Age | 5-year age group categorization with outlier handling; top-coding at 80 years and above |
|  | Household Size | Recategorized into five groups: 1, 2, 3, 4, and 5 or more members |
| General Information | Date | Provided at the daily level with added noise ($\pm 5$ days) |
|  | Drug Information | Last three digits masked |
| Sensitive Information | Disease Code | 493 sensitive disease codes suppressed or masked |

## RESULT

### 1. Annual Application and Approval Trends

A total of 311 research projects were submitted through the HCDL from 2022 to 2025, of which 190 (61.1%) were approved (Figure 4). The annual number of applications increased steadily, from 44 in 2022 to 75 in 2023, 87 in 2024, and 105 in 2025—representing an approximately 2.4-fold increase over the four-year period. The number of approved projects also reflects an overall upward trend from 29 in 2022 to 66 in 2025. Annual approval rates were 65.9%, 66.7%, 51.7% (2024), and 62.9% (2025), remaining within a range of approximately 50% to 67% across the study period.

**Figure 4.**
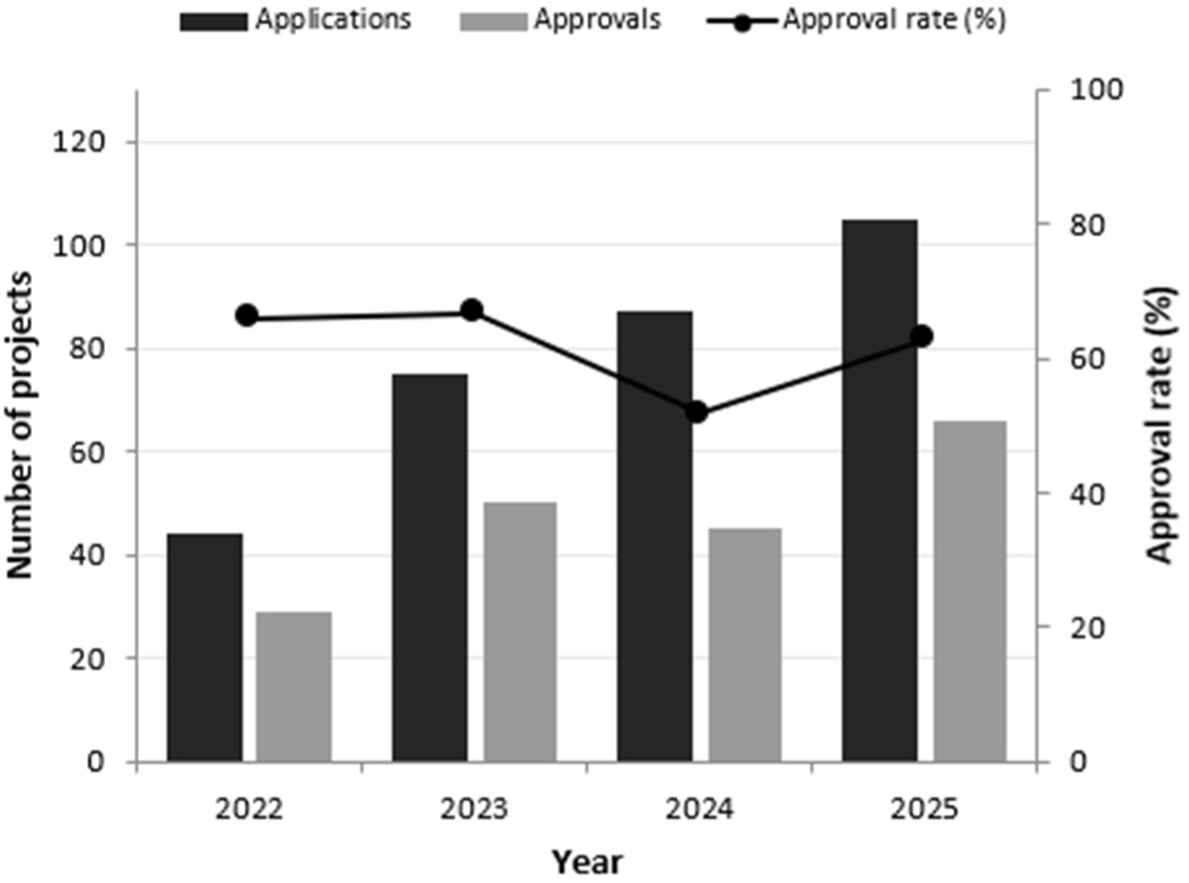
Annual application and approval trends. Black bars represent the annual number of applications from 2022 to 2025, and grey bars represent the annual number of approved projects over the same period. Dots indicate the annual approval rate.

### 2. Linkage Rates with Reference Datasets

To examine whether the platform’s TTP-based linkage method secures sample sizes sufficient for research, linkage rates with major databases were calculated for two reference datasets with distinct population characteristics (Table 3). Here, the linkage rate refers to the proportion of subjects in the reference dataset who were present in and linked to a given database. For both KNHANES (total n = 117,784) and DON (total n = 40,179), linkage rates with NHIS SCREEN (eligibility) and HIRA CLAIMS were generally high, at around 80% or above. This confirms that the platform can reliably secure sample sizes adequate for research even though it does not exchange direct personal identifiers during linkage. By contrast, linkage rates with mortality data (MORT) were considerably lower, at 5.2% and 14.6%, respectively, reflecting the nature of mortality records, which are limited to individuals who died during the observation period.

**Table 3.** Linkage rates between reference datasets and other institutional databases.

| Reference dataset (n) | Linked database | Linkage rate (%) | Linked records (n) |
| --- | --- | --- | --- |
| KNHANES<br>(117,784) | SCREEN (eligibility) | 84.7 | 99,781 |
|  | CLAIMS | 83.8 | 98,706 |
|  | MORT | 5.2 | 6,063 |
| DON<br>(40,179) | SCREEN (eligibility) | 85.8 | 34,509 |
|  | CLAIMS | 77.5 | 31,134 |
|  | MORT | 14.6 | 5,865 |

### 3. Database Use

Analysis of the databases used across the 190 approved research projects revealed that CLAIMS was requested in 177 projects (93.2%), representing the highest demand, followed by SCREEN in 156 projects (82.1%) (Figure 5A). MORT was requested in 96 projects (50.5%), KNHANES in 92 (48.4%), and CANCER in 60 (31.6%), constituting a second tier of frequently used databases. In contrast, the IMM (21), DON (19), DEM (10), AIR (8), TUBER (5), MIL (5), ILSAN (1), and KoGES (0) databases exhibited comparatively low use rates. Analysis of the number of databases requested per project showed that projects linking data from four institutions were the most common with 82 projects (43.2%), followed by three institutions (70 projects, 36.8%), two institutions (26 projects, 13.7%), and five institutions (12 projects, 6.3%) (Figure 5B). Projects linking data from three or more institutions accounted for 86.3% of the total.

**Figure 5.**
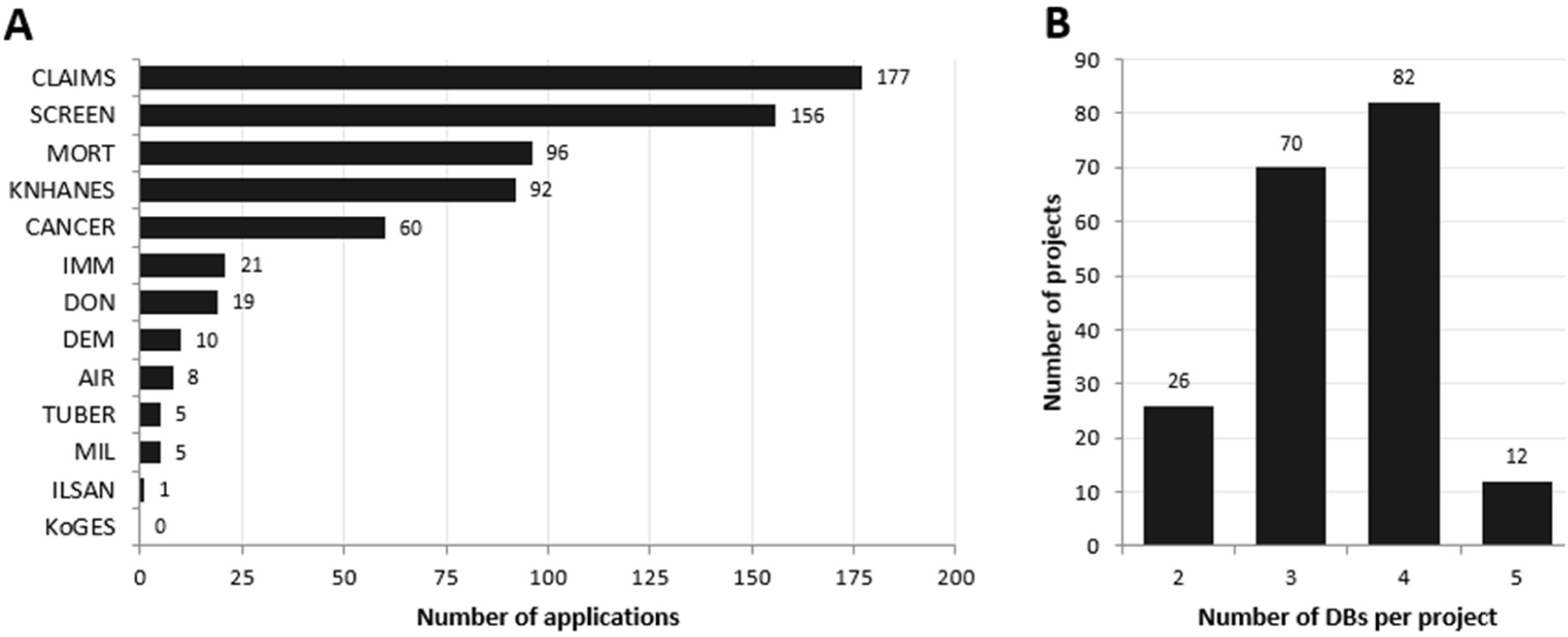
Database Use. **A.** Number of applications by database among the 190 approved projects. **B.** Number of databases requested per project

### 4. Database Linkage patterns

Analysis of database linkage patterns revealed that the SCREEN-CLAIMS-KNHANES combination was the most frequently used (n = 22), followed by SCREEN-CLAIMS-KNHANES-MORT (n = 21) and SCREEN-CLAIMS-CANCER-MORT (n = 19) (Figure 6). CLAIMS and SCREEN served as core datasets in most linkage, while KNHANES, MORT, and CANCER played complementary roles, contributing information on lifestyle factors, vital status, and cancer diagnosis, respectively.

**Figure 6.**
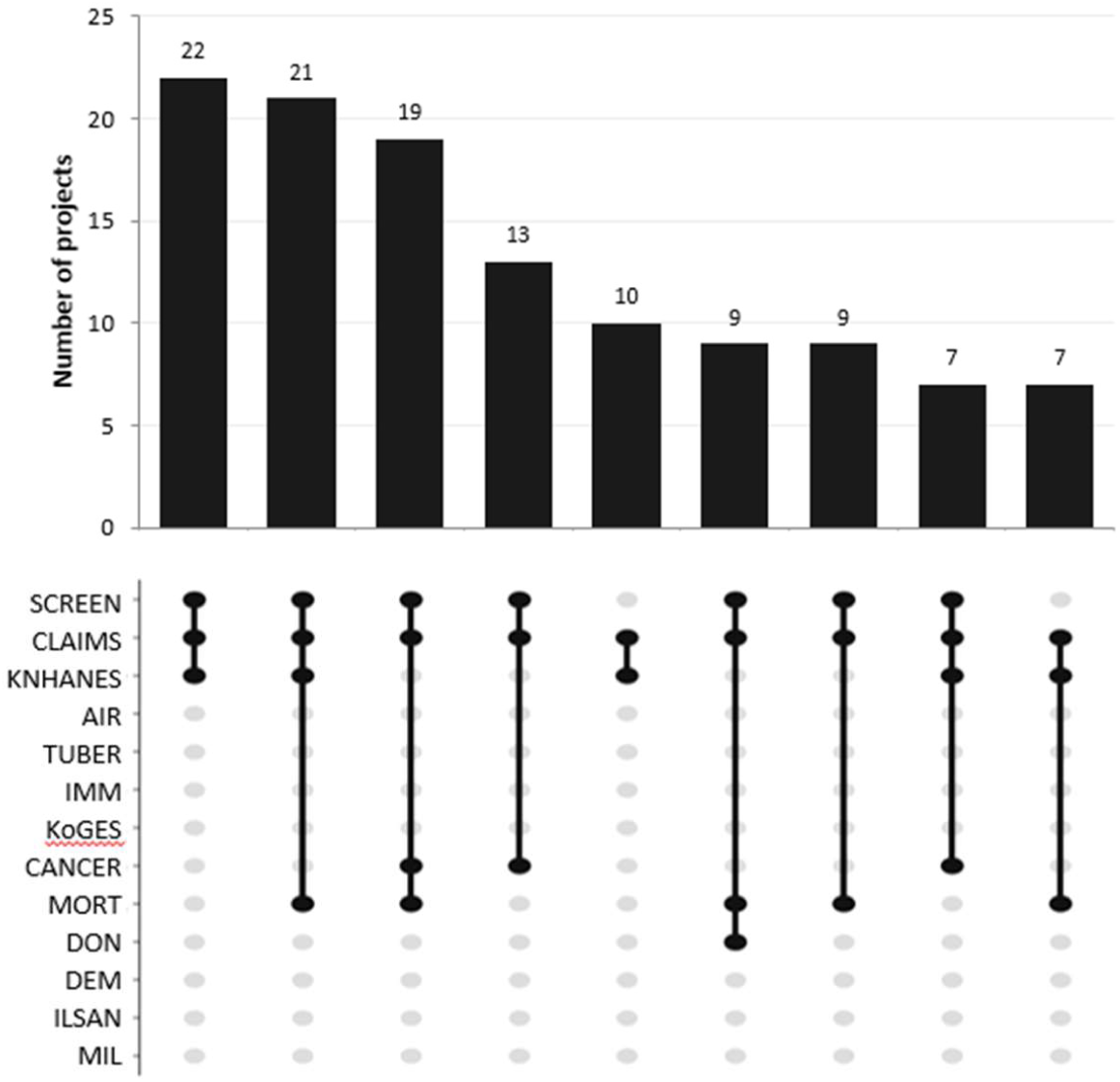
Co-occurrence combination patterns of databases. *Top*. The bar chart shows the number of projects for each database combination in descending order. *Bottom*. Black dots connected by a vertical line represent a single co-occurrence combination. Only the nine most frequent combinations are shown.

### 5. Co-occurrence pattern of databases

The linkage relationships among databases across the 190 approved research projects were visualized as a network graph (Figure 7). Node size reflects the frequency of database use while line thickness indicates the frequency with which two databases co-occurred within the same project. The distance between nodes is inversely proportional to co-occurrence frequency, such that databases frequently used together are positioned closer together.

**Figure 7.**
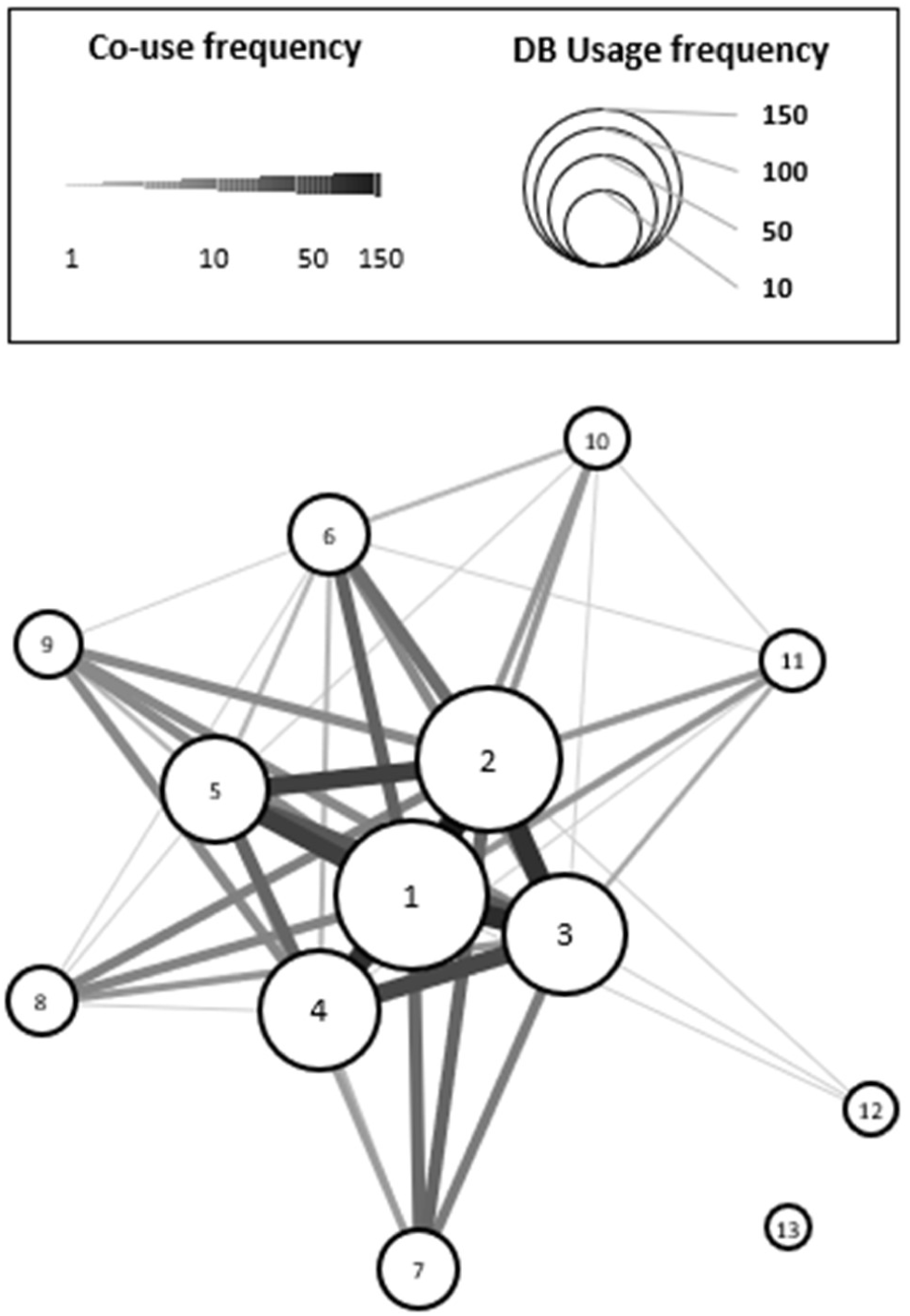
Database co-occurrence network. Each node represents a database, with node size proportional to its usage frequency shown in Figure 5A. Lines connect two databases that were used together in a project, with line thickness proportional to co-occurrence frequency. Node numbers correspond to the following databases: *1*, CLAIMS; *2*, SCREEN; *3*, MORT; *4*, KNHANES; *5*, CANCER; *6*, IMM; *7*, DON; *8*, DEM; *9*, AIR; *10*, TUBER; *11*, MIL; *12*, ILSAN; *13*, KoGES. These identifiers are also listed in Table 4.

**Table 4.**
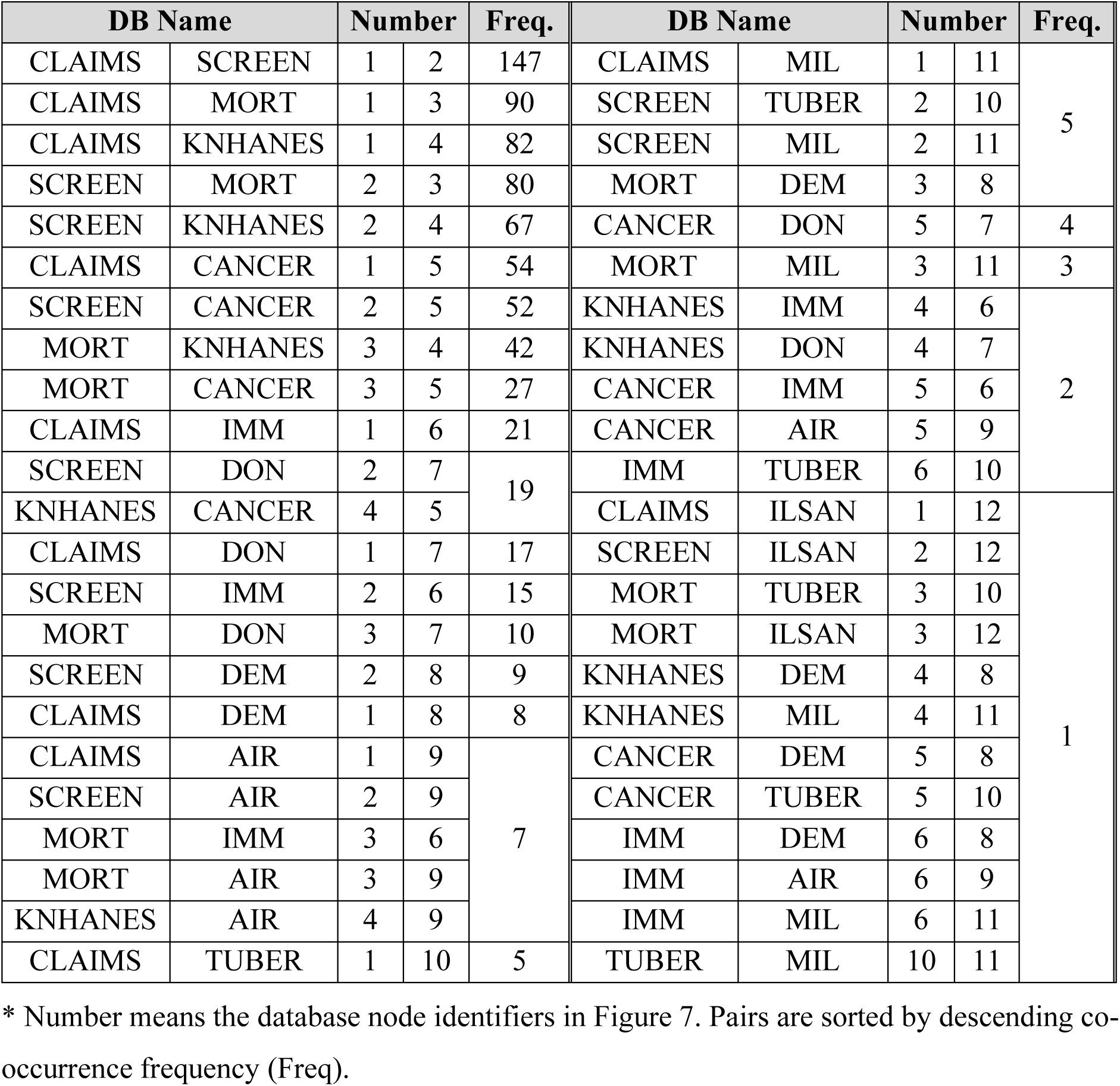
Co-occurrence frequency of database pairs.

SCREEN and CLAIMS were used together in 147 projects (77.4 %), representing the highest co-occurrence frequency among all database pairs, and are positioned adjacently at the center of the network. Both databases co-occurred with the majority of other databases (SCREEN with 11, CLAIMS with 10 out of 13), confirming their role as central backbone databases within the network. The next most frequent pairings were CLAIMS–MORT (n = 90), CLAIMS–KNHANES (n = 82), SCREEN–MORT (n = 80), and SCREEN–KNHANES (n = 67), indicating that MORT and KNHANES are strongly connected to the central SCREEN–CLAIMS cluster. CANCER also showed strong connections with CLAIMS, SCREEN, and MORT (n = 54, 52, 27, respectively), forming part of the core network alongside SCREEN, CLAIMS, MORT, and KNHANES. In contrast, KoGES (n = 0), ILSAN (n =1), and MIL, TUBER, and AIR (n =2 each) were located at the periphery of the network, reflecting their relatively limited co-occurrence with other databases.

## DISCUSSION

This study examined the structural design of the Healthcare Big Data Linkage Platform (HCDL) and analyzed data use trends from 2022 to 2025. The platform was established to overcome the fragmented data access process in which researchers were required to contact multiple data-providing institutions and obtain separate approvals. It provides a unified platform for secure data access, linkage, and analysis of multiple healthcare databases within a single, integrated system. It centralizes multi-institutional review within a single governance framework, thereby reducing administrative burden. This also enables research designs that were previously impractical by allowing flexible combinations of public datasets.

### Sustained Growth in Data-Driven Research Demand

The steady year-over-year increase in data applications to the HCDL from 2022 to 2025 provides empirical evidence of rapidly growing demand for data linkage in healthcare research. This trend is consistent with the global expansion of data-driven research and increasing emphasis on precision medicine (Jung et al., 2024), suggesting that the platform is becoming a key infrastructure for supporting this growing demand.

Approval rates remained relatively stable throughout the period, ranging from 51.7% to 66.7%, despite a more than twofold increase in application volume. This suggests that the review system has maintained consistent evaluation standards. However, an overall approval rate of approximately 60% points to two potential issues. First, it suggests that the current review process could be further optimized. Second, it may reflect the inclusion of applications with underdeveloped research designs or insufficiently specified data use plans. Given that the Korea Health Information Service (KHIS) currently offers annual training on healthcare data use, combining pre-application support with these programs may help improve both approval rates and overall research quality.

### The Centrality of HIRA and NHIS

The markedly high use of CLAIMS (177 projects; 93.2%) and SCREEN (156 projects; 82.1%) indicates that these databases function as the primary foundational resources for studies that integrate data across multiple institutions within the platform (Figure 5). Administrative claims data within the CLAIMS comprehensively capture healthcare use across the national population and provide the basis for defining study cohorts and assessing clinical outcomes. (Rho et al., 2021). In parallel, SCREEN include eligibility information, insurance premium records, and health screening data, and are widely used to characterize the socioeconomic status and baseline health conditions of study populations. The high utilization rates of CLAIMS and SCREEN on the platform reflect researchers’ continued reliance on real-world evidence resources representative of large-scale clinical practice. Korea’s single-payer nationwide claims data offer high statistical power (Choi, 2020) and, given their complete enumeration survey design, provide an analytical basis for investigating rare diseases and infrequent adverse events that would otherwise remain difficult to detect through conventional clinical trials alone (Ryu, 2017). These characteristics constitute the academic rationale for the position of CLAIMS and SCREEN as the central datasets on the platform.

This centrality is further supported by the co-occurrence network (Figure 7), where CLAIMS and SCREEN exhibit the highest degree of connectivity among all databases. The network structure, in which MORT and KNHANES are closely linked to the SCREEN–CLAIMS core, suggests that integrating healthcare use data with mortality, lifestyle, and nutritional information represents a dominant research paradigm. In addition, the linkage of CANCER to the SCREEN–CLAIMS core reflects strong demand for combining detailed cancer data with healthcare insurance records to examine treatment patterns, outcomes, and long-term trajectories in cancer populations.

In contrast, databases focused on specific diseases or population subgroups, such as KoGES, ILSAN, and TUBER, are accessed less frequently. Although this partly reflects their relevance to more specialized research topics, it may also suggest limited awareness of their research value. These findings indicate that more active dissemination and promotion may be needed to enhance their use.

### The Trend Toward Integrative Research

The tendency in the use of an average of three or more databases per project reflects increasing demand for understanding complex health outcomes that cannot be adequately explained by a single data source (Lee et al., 2024). Combining healthcare data from HIRA and NHIS with lifestyle and nutritional information from KNHANES, information of the deceased from the MORT database, and cancer registry from CANCER enables longitudinal analyses spanning the full disease trajectory, from onset and progression to outcomes. K-CURE, the national cancer data initiative, is a representative example, integrating cancer registry, screening, claims, and mortality data with clinical data from multiple medical institutions to support full-cycle analysis of disease (Choi et al., 2024; Lee et al., 2025). This pattern represents a shift toward more comprehensive and integrative approaches for healthcare research, and suggests that the platform is supporting this evolving research trend.

The frequent observation in co-occurrence pattern analyses (Fig. 7) of CLAIMS and SCREEN being linked with MORT and KNHANES suggests that cohort studies tracking prognosis by linking clinical events with mortality outcomes constitute the predominant research form. This pattern is expected to be particularly pronounced in research domains requiring long-term follow-up, such as chronic diseases, cancer, and geriatric health.

### Strengths and Limitations of the TTP-Based Linkage Method

The TTP-based linkage method adopted by the platform structurally reduces the risk of exposing personal information, as institutions link data through keys issued by a trusted third party rather than exchanging direct identifiers such as resident registration numbers. The linkage rate analysis demonstrates that this privacy-preserving method nonetheless secures sample sizes usable for research: across two reference datasets with distinct population characteristics, linkage rates with SCREEN (eligibility) and CLAIMS were consistently around 80% (Table 3), confirming that a pseudonymized approach can reliably yield analytic populations sufficient for a wide range of studies. However, because indirect identifier-based linkage inevitably entails a certain proportion of missed matches (Lee et al., 2024), it can weaken statistical power and biased estimates in studies with small populations, such as those involving rare diseases or specific subgroups. This approach may therefore be unsuitable for studies in which the target population is expected to be small. Previous studies have suggested the potential to improve linkage accuracy to over 90% by introducing a hybrid linkage methodology combining probabilistic and deterministic linkage (Batra et al., 2024), and has demonstrated genuine privacy-preserving analysis techniques based on multiparty homomorphic encryption (Froelicher et al., 2021). Applying these methods could simultaneously ensure the dual demands of maintaining linkage accuracy and protecting personal information.

### Equity of Access and Significance of the Platform as Research Infrastructure

The study demonstrates that the HCDL has facilitated the expansion of researches linking multiple data sources by addressing the limitation of previously fragmented data access systems. The 2.4-fold increase in applications over four years reflects the practical benefits of the platform’s unified governance structure and streamlined data access procedures. Under the conventional approach through dedicated linkage agencies, delays or coordination failures at a single data-holding institution could disrupt the entire research process. In contrast, the previously time-consuming data acquisition process, including institution-by-institution review and linkage queues, can now be completed within approximately six months through a single application. This represents a substantial improvement in both research efficiency and the predictability of the data acquisition process. Furthermore, the HCDL also addresses structural inequities in data access in healthcare research. Under the previous system, access to data was largely determined by a researcher’s institutional affiliation, leading to disparities based on organizational capacity. By enabling researchers to apply for and use data from participating public agencies on equal terms regardless of institutional affiliation, the HCDL has enhanced equity in research opportunities and expanded the diversity of the healthcare research ecosystem. The equity is not limited to the institutional level, but extends to the regional level: the SDCs distributed across regions nationwide allow researchers to access data analysis environments irrespective of their physical location. Moreover, the SDCs operate as closed analysis environments, thereby balancing the dual demands of expanded accessibility and secure use. The fact that the platform has provided data to 190 research projects over four years under a stable governance framework empirically supports its establishment as a core infrastructure for health data research in South Korea.

### Future Directions

To further consolidate its role as a core infrastructure for healthcare research, several aspects should be addressed. The number of data extraction personnel, virtual analysis accounts, and SDCs should be expanded to meet growing demand. Second, data linkage could be extended beyond claims data to include clinical sources such as electronic medical records (EMRs), thereby enabling a wider range of research applications (Jung et al., 2024). Third, the development of a standardized metadata framework would strengthen interoperability with other systems. Recently, HIRA claims data that covers approximately 56.4 million individuals over a ten-year period was converted to an analysis-ready OMOP-CDM research resource (Yu et al., 2025). This initiative provides a concrete technical roadmap for future expansion toward internationally compatible analytical infrastructure. Fourth, active guidance and training programs on the characteristics of each database and their applications in research design should be provided, thereby promoting wider use of the diverse databases, including those that are currently underused. Taken together, these improvements would support the platform’s role as a key infrastructure for evidence-based health policy and precision medicine research.

## ABBREVIATION

CANCER: Cancer Registry DB from NCC
DEM: Dementia DB from NMC
DON: Donor and Recipient Information DB from NIOTBM
DRB: Data Review Board
HCDL: Healthcare Big Data Linkage Platform
HIRA: Health Insurance Review and Assessment Service
ILSAN: Patient Information DB from Ilsan Hospital
IMM: National Immunization DB from KDCA
IRB: Institutional Review Board
KDCA: Korea Disease Control and Prevention Agency
KHIS: Korea Health Information Service
KNHANES: Korea National Health and Nutrition Examination Survey DB from KDCA
KoGES: Korean Genome and Epidemiology Study DB from KDCA
MIL: Physical Examination and Military Draft Screening Results DB from MMA
MMA: Military Manpower Administration
MODS: Ministry of Data and Statistics
MOHW: Ministry of Health and Welfare
MORT: Cause of Death Statistics DB from MODS
NCC: National Cancer Center
NECA: National Evidence-based Healthcare Collaborating Agency
NHIS: National Health Insurance Service
NIOTBM: National Institute of Organ, Tissue and Blood Management
NMC: National Medical Center
NRC: National Rehabilitation Center
POL: KNHANES Air Pollution DB
REC: Research Evaluation Committee
REHAB: Nursing Observation Records and Medical Record Information DB from NRC
SDC: Secure Data Center
TTP: Trusted Third Party

## ETHICS APPROVAL

This study did not require Institutional Review Board approval, as it analyzed only aggregate administrative records of the platform (application and approval statistics) and did not involve any individual-level patient data or human subjects.

## DATA AVAILABILITY STATEMENT

All data produced in the present study are available upon reasonable request to the authors.

## REFERENCE

Batra, K., Goel, V., Reyes, A., Assoumou, B., Simangan, D. P., Abdulla, F., & Kuhls, D. A. (2024). Unifying and linking data sources in medical and public health research. Journal of Medicine Surgery and Public Health, 5, 100164–100164. 10.1016/j.glmedi.2024.100164

Choi, E.-K. (2020). Cardiovascular Research Using the Korean National Health Information Database. Korean Circulation Journal, 50, 663–672. 10.4070/kcj.2020.0171

Choi, D., Guk, M. Y., Kim, H. R., Ryu, K. S., Kong, H., Soung, H., Kim, H., Chae, H., Jeon, Y. S., Kim, H., Jung, J., Im, J.-S., & Choi, K. S. (2024). Data Resource Profile: The Cancer Public Library Database in South Korea. Cancer Research and Treatment, 56(4), 1014–1026. 10.4143/crt.2024.207

Froelicher, D., Troncoso-Pastoriza, J. R., Raisaro, J. L., Cuendet, M. A., Sousa, J. S., Cho, H., Berger, B., Fellay, J., & Hubaux, J. (2021). Truly privacy-preserving federated analytics for precision medicine with multiparty homomorphic encryption. Nature Communications, 12(1), 5910–5910. 10.1038/s41467-021-25972-y

Herrett, E., Gallagher, A. M., Bhaskaran, K., Forbes, H., Mathur, R., Staa, T. van, & Smeeth, L. (2015). Data Resource Profile: Clinical Practice Research Datalink (CPRD). International Journal of Epidemiology, 44(3), 827–836. 10.1093/ije/dyv098

Jang, Y., Lee, H., & Park, H.-K. (2025). Surveillance System for Infectious Disease Prevention and Management: Direction of Korea’s Infectious Disease Surveillance System. Journal of Korean Medical Science, 40(8). 10.3346/jkms.2025.40.e108

Jung, J., Jhang, H., Park, K., & Yun, J. E. (2024). A Survey on the Utilization of Big Data in the Healthcare Sector and the Demand for Integrated Data. Journal of Health Informatics and Statistics, 49(1), 62–69. 10.21032/jhis.2024.49.1.62

Kang, C., Lee, H., Han, J., Kim, J., & Lee, Y. (2018). Types of Secondary Data Sources and Their Utilization for Health and Medical Status Analysis. Gyeonggi-do Public Health Policy Institute.

Kim, J.-A., Yoon, S., Kim, L. Y., & Kim, D. (2017). Towards Actualizing the Value Potential of Korea Health Insurance Review and Assessment (HIRA) Data as a Resource for Health Research: Strengths, Limitations, Applications, and Strategies for Optimal Use of HIRA Data. Journal of Korean Medical Science, 32(5), 718–718. 10.3346/jkms.2017.32.5.718

Lee, B., Lee, Y., Kim, S., Oh, H., Won, S., Jang, S., Jeon, Y. J., Yoo, B., & Bak, J.-K. (2024). Impact of linkage level on inferences from big data analyses in health and medical research: an empirical study. BMC Medical Informatics and Decision Making, 24(1), 193–193. 10.1186/s12911-024-02586-0

Lee, S., Choi, Y. H., Kim, H. M., Hong, M., Park, P., Kwak, I. H., Kang, Y. J., Choi, K. S., Kong, H., Cha, H., Kim, H., Ryu, K. S., Jeon, Y. S., Kim, H., Jung, J. M., Im, J.-S., & Chae, H. (2025). The Cancer Clinical Library Database (CCLD) from the Korea-Clinical Data Utilization Network for Research Excellence (K-CURE) Project. Cancer Research and Treatment, 57(1), 19–27. 10.4143/crt.2024.218

Leitsalu, L., Alavere, H., Tammesoo, M., Leego, E., & Metspalu, A. (2015). Linking a Population Biobank with National Health Registries—The Estonian Experience. Journal of Personalized Medicine, 5(2), 96–106. 10.3390/jpm5020096

Musa, S. M., Haruna, U. A., Manirambona, E., Eshun, G., Ahmad, D. M., Dada, D. A., Gololo, A. A., Musa, S. S., Abdulkadir, A. K., & Lucero-Prisno, D. E. (2023). Paucity of Health Data in Africa: An Obstacle to Digital Health Implementation and Evidence-Based Practice. Public Health Reviews, 44, 1605821–1605821. 10.3389/phrs.2023.1605821

Oh, K., Kim, Y., Kweon, S., Kim, S., Yun, S., Park, S., Lee, Y.-K., Kim, Y., Park, O., & Jeong, E. K. (2021). Korea National Health and Nutrition Examination Survey, 20th anniversary: accomplishments and future directions. Epidemiology and Health, 43. 10.4178/epih.e2021025

Park, S., & Cho, E. (2014). National Infectious Diseases Surveillance data of South Korea. Epidemiology and Health, 36. 10.4178/epih/e2014030

Rho, Y., Cho, D. Y., Son, Y., Lee, Y. J., Kim, J., Lee, H. J., You, S. C., Park, R. W., & Lee, J. Y. (2021). COVID-19 International Collaborative Research by the Health Insurance Review and Assessment Service Using Its Nationwide Real-world Data: Database, Outcomes, and Implications. Journal of Preventive Medicine and Public Health, 54(1), 8–16. 10.3961/jpmph.20.616

Ryu, D.-R. (2017). Introduction to the Medical Research Using National Health Insurance Claims Database. Korean Journal of Internal Medicine, 32, 973–976. 10.3904/kjim.2017.284

Vilpponen, H., Piirainen, A., Kallberg, M., & Mikkonen, T. (2024). Secondary Use of Health Data: Centralized Structure and Information Security Frameworks in Finland. In arXiv (Cornell University). Cornell University. 10.48550/arxiv.2412.06800

Yu, D. H., Lim, S., Shin, H., Kim, M., You, S. C., Park, R. W., & Kim, C. (2025). Data Resource Profile: Health Insurance Review and Assessment Service Korean Nationwide Claims OMOP-CDM (2015–2024) database, "HIRA K-OMOP". medRxiv 2025.12.08.25341603. 10.64898/2025.12.08.25341603

